# Differential clinical characteristics across traditional Chinese medicine (TCM) syndromes in patients with sickle cell disease

**DOI:** 10.1101/2023.10.08.23296714

**Authors:** Ying Wang, David D Wang, Andrew Q Pucka, Andrew RW O’Brien, Steven E Harte, Richard E Harris

**Affiliations:** Department of Anesthesia, Stark Neurosciences Research Institute, Indiana University School of Medicine, Indianapolis, IN, USA; Division of Hematology/Oncology, Department of Medicine, Indiana University School of Medicine, Indianapolis, IN, USA; Center for Integrative Health, The Ohio State University, Columbus, OH, USA; Indiana University Simon Cancer Center, Indianapolis, IN, USA; Chronic Pain and Fatigue Research Center, Department of Anesthesiology, University of Michigan Medical School, Ann Arbor, MI, USA; Susan Samueli Integrative Health Institute, and Department of Anesthesiology and Perioperative Care, School of Medicine, University of California at Irvine, Irvine, CA, USA

**Keywords:** Sickle cell disease, Pain, Traditional Chinese Medicine, Syndrome differentiation, Acupuncture, Patient-reported outcomes, Quantitative sensory testing, Morphine milligram equivalents

## Abstract

**Background:** Pain is a common, debilitating, and poorly understood complication of sickle cell disease (SCD). The need for clinical pain management of SCD is largely unmet and relies on opioids as the main therapeutic option, which leads to a decreased quality of life (QoL). According to the literature, acupuncture has shown certain therapeutic effects for pain management in SCD. However, these clinical studies lack the guidance of Traditional Chinese Medicine (TCM) Syndrome Differentiation principles for treatment.

**Aim:** To characterize differences in clinical presentation amongst TCM-diagnosed syndromes in SCD patients.

**Method:** 52 patients with SCD and 28 age- and sex-matched healthy controls (HCs) were enrolled in an ongoing trial of acupuncture. Each participant completed a series of questionnaires on pain, physical function, fatigue, sleep, anxiety, depression, and QoL and underwent cold- and pressure-based quantitative sensory testing at baseline. Data on prescription opioid use over the 12 months prior to study enrollment was used to calculate mean daily morphine milligram equivalents (MME). Differences among the three TCM syndromes were analyzed by one-way ANOVA followed by Tukey post hoc testing. Two-sample t-tests were used to compare SCD and HC groups.

**Results:** TCM diagnosis criteria classified SCD patients into one of three TCM syndromes: a) Equal; b) Deficiency; and c) Stagnation. The Stagnation group exhibited higher pain interference, physical dysfunction, nociplastic pain, fatigue, anxiety, depression, MME consumption, and lower sleep quality and QoL compared to the Equal group. Few differences were observed between HCs and the Equal SCD group across outcomes. Deficiency and Stagnation groups were differentiated with observed- and patient-reported clinical manifestations.

**Conclusion:** These findings suggest that TCM-diagnosed syndromes in SCD can be differentially characterized using validated objective and patient-reported outcomes. Because characteristics of pain and co-morbidities in each SCD patient are unique, targeting specific TCM “syndromes” may facilitate treatment effectiveness with a syndrome-based personalized treatment plan that conforms to TCM principles. These findings lay the foundation for the development of tailored acupuncture interventions based on TCM syndromes for managing pain in SCD. Larger samples are required to further refine and validate TCM diagnostic criteria for SCD.

## 1. Introduction

Pain in sickle cell disease (SCD) is both common and debilitating. While the pain can be chronic in nature, severe acute vaso-occlusive pain crises (VOC) are also present in many of these individuals, resulting in recurrent pain episodes that are unpredictable, and frequently require hospitalization. As such VOCs are a leading factor lowering the quality of life (QoL) in SCD (1). While opioids are the main therapeutic option for managing these events, there are limited alternative and effective treatments beyond opioids for these patients (2).

Acupuncture, a mainline component of traditional Chinese medicine (TCM), has been used for over four-thousand years and has been recognized as an effective treatment approach for chronic and acute pain conditions (3, 4). Emerging data suggests that acupuncture may alleviate pain in both adult and pediatric patients with SCD (5–8). However, a key principle of TCM practice is “treatment based on differentiation of syndrome” wherein patients are evaluated on a set of diagnostic criteria which can allow classification into a specific TCM syndrome for customized treatment. This classification, along with consideration of additional individual specific symptom patterns, guides the tailored selection of acupuncture points to treat a particular patient (9, 10).

Two recent studies categorized different TCM patterns in women with pelvic pain (11) or fibromyalgia (12). These studies showed that there were 6 TCM patterns in female patients with pelvic pain whereas 3 TCM patterns in women with fibromyalgia. There have been no studies on TCM syndrome differentiation in SCD. The small sample size and failure to apply TCM principles in previous studies of SCD have made it difficult to assess the full potential benefit of acupuncture for SCD. For example, it is not known if subgroups of SCD patients exhibit different pathogenic features when classified by TCM principles. Identification of different subtypes may also be relevant for other therapies in SCD.

The classic pathophysiology of SCD is polymerization of deoxy sickle hemoglobin which leads to a complex interplay of disrupted red blood cell rheology, hemolysis, inflammation, and vasculopathy. However, pain in SCD is not well understood, and is characterized by substantial heterogeneity in clinical manifestation and underlying pathophysiology (16–24). Notably, clinical pain varies in many aspects including quality, intensity, location, and duration under both chronic and VOC stages. The onset of VOCs can happen anywhere in the body, with the most painful sites being consistent or fixed, and ranging from mild to severe intensity during a specific VOC crisis. The frequency of VOCs also varies widely in SCD with a recent systematic review reporting a range of 0 to 18.2 per year (25), and variable distribution of VOCs throughout the year between individuals. Finally, occurrence of VOCs can also be triggered by various multidimensional factor(s), such as temperature, mental stress, dehydration, and/or fatigue. These individual differences add significant challenges in the currently unmet management of clinical pain in SCD and thus strongly warrants the need for individualized diagnosis and personalized treatment. Establishing TCM syndrome differentiation in SCD to help guide acupuncture treatment is the current priority.

Here we report preliminary baseline data from our ongoing randomized sham-controlled trial of acupuncture for SCD (ClinicalTrials.gov Identifier: NCT05045820). Patients with SCD were classified into three different TCM syndrome groups prior to treatment according to TCM principles. Our objective was to investigate differences in patient reported outcome measures (PROMs), quantitative sensory testing (QST), and blood-based assessments across these TCM syndrome groups.

## 2. Methods

### 2.1. Participants

Fifty-two patients with SCD (25 male and 27 female), aged 14-73, were enrolled. The main inclusion eligibility criteria included: 1) experiencing chronic pain in the past 6 months or at least one VOC in the past 12 months, 2) no recent history of initiating or adjusting the dose of stimulant medications, 3) willing to maintain current treatments, and 4) willing to not introduce any new medications or treatment modalities for control of pain symptoms during the study. Detailed inclusion and exclusion criteria can be found in Supplementary Table 1. Participants’ demographics can be found in Table 1. Twenty-eight pain-free ethnicity-, age- and gender-matched participants without SCD were enrolled as healthy controls (HCs). A subset of SCD and HCs were matched on age and gender for comparisons of PROMs (n=25-27 each) and QST (n=21-28 each).

**Table 1.** Demographics. Demographic information of participants in each TCM group and HCs were collected during the screening visit. The demographic indexes did not differ across groups except that weight and body mass index was significantly lower in Deficiency compared to HCs and Stagnation.

| Subjects' characteristics (n=52) | TCM#1 (Equal)<br>(n=13) | TCM#2 (Deficiency)<br>(n=20) | TCM#3 (Stagnation)<br>(n=19) | Healthy<br>(n=28) |
| --- | --- | --- | --- | --- |
| Age (year), median (range) | 37 (14-49) | 36.5 (18-73) | 30 (14-58) | 33 (14-65) |
| Female, n (%) | 5 (38.5) | 9 (45.0) | 12 (63.2) | 17 (60.7) |
| Height (cm), median (range) | 173.4 (161.5 - 185.2) | 168.9 (150.4-184.3) | 168.1 (154.7 - 181.1) | 168.15 (154.8 - 184.4) |
| Weight (kg), median (range) | 73.4 (54.4 - 112.6) | 62.9 (37.6-106.2) **## | 80.8 (56.8 - 123.5) | 75.8 (50.4 - 112.4) |
| Body mass index, median (range) | 23.7 (19.3 - 33.7) | 20.6 (16.5 - 35.6) **### | 28.6 (19.7 - 35.6) | 26.7 (19 - 42.8) |
| **SCD type diagnosis (n=52)** |  |  |  |  |
| SS or Sβ0/SC/Sβ+ thalassemia<br>(n / n / n) | 7 / 6 / 0 | 15 / 1 / 4 | 11 / 7 / 1 | N/A |
| **Hematological treatment (n=52)** |  |  |  |  |
| Chronic transfusion therapy, n<br>(%) | 0 | 3 (15) | 1 (7) | N/A |
| Hydroxyurea, n (%) | 7 (53.8) | 14 (70) | 8 (53.8) | N/A |

\*\*p< 0.01 compared with Healthy, ##p< 0.01 compared with Stagnation, ###p< 0.001 compared with Stagnation

Patients underwent a TCM diagnostic examination and phenotyping with validated PROMs, QST, and blood tests. HCs completed PROMs, QST, and blood tests. The study was approved by the Institutional Review Board at Indiana University. All patients provided written informed consent before the study.

### 2.2. TCM diagnosis

Participants who completed the baseline stage of data collection were then randomized to either verum or sham acupuncture for 5 weeks. All data presented here are cross-sectional at baseline. Longitudinal results related with acupuncture intervention will be reported elsewhere. TCM diagnosis was performed before the treatment by one practitioner with over 15 years of clinical experience. This practitioner utilized a standardized procedure with TCM diagnostic methods such as “Observation,” “Listening and Smelling,” “Inquiry,” and “Pulse-feeling and palpation” (see Supplementary Table 2) (26). Following differentiation of syndrome, patients were classified into one of three TCM syndrome groups: a) “Stagnation greater than deficiency” (abbreviated as “Stagnation”), b) “Deficiency greater than stagnation” (abbreviated as “Deficiency”), and c) “Equal stagnation and deficiency” (abbreviated as “Equal”). Diagnoses were confirmed in a blinded manner by a second practitioner with more than 20 years of experience.

### 2.3. Patient reported outcome measures (PROMs)

Pain (pain intensity and interference), physical function, and satisfaction with one’s social role were assessed with the PROMIS-29 (27). Neuropathic pain symptoms were evaluated using the PainDETECT (28, 29). The Fibromyalgia Survey Questionnaire (FSQ), which consists of the Widespread Pain Index and the Symptom Severity scale (30, 31), was utilized as a surrogate measure of nociplastic pain. Fatigue and quality of sleep were assessed using the Multi-dimensional Fatigue Inventory (MFI) (32) and the Pittsburgh Sleep Quality Index (PSQI) (17), respectively. Anxiety and depression were evaluated using Hospital Anxiety and Depression Scale (HADS) (17).

### 2.4. Quantitative sensory testing (QST)

QST is an established experimental protocol that examines ascending excitatory and descending inhibitory aspects of pain processing by assessing an individual’s perceptual response to applied stimuli. QST was conducted at up to three different body sites: the primary testing site was the most painful area of the body as reported by each patient; the dominant-side ventral forearm, and/or the dominant-side upper trapezius muscle as in previous SCD studies (33, 34). For non-SCD controls, primary testing sites matched those identified in SCD patients in addition to testing at the dominant forearm and/or trapezius.

#### Thermal (heat/cold) Detection / Pain Threshold

was determined at each testing site using a TCA11 (QST-Lab, France) with a thermal probe in contact with the subject’s skin surface. The thermode temperature was increased/decreased from a baseline temperature by adapting to the individual ‘s own body temperature at a rate of 0.5-1 °C/s. Subjects indicated the thermal detection threshold (when they first felt the thermal stimuli) and the pain threshold (when they first felt pain from the thermal stimuli). The average of three trials of each test was used for analysis.

#### Mechanical Detection Threshold / Mechanical Pain Threshold

was examined using von Frey monofilaments (Stoelting Co., USA) and calibrated pinprick stimuli (MRC Systems GmbH, Germany). Each von Frey monofilament was applied three times in ascending sequence until the stimulus was detected in at least two of the three trials. Then, the next lower von Frey monofilament was applied, and the lowest filament to be detected at least twice was determined as the mechanical detection threshold. The mechanical pain threshold was determined using different pinprick probes applied to the skin surface of each site. Testing started with a stimulation intensity of 8 mN and in each case, the next higher pinprick stimulator was applied until the perception of “touch” changed its quality towards an additional “sharp”, “pricking” or “stinging” impression. The corresponding intensity represented the first suprathreshold value. Once the first painful stimulus was perceived, the testing direction was changed towards lower stimulus intensities until the first stimulus, when applied to the skin, that was perceived as “blunt” and no longer as being “sharp”, “pricking” or “stinging” (subthreshold value). Again, a directional change towards higher intensities occurred and the cycle was repeated until all five supra- and five subthreshold values were found that represent the inflection point to determine the mechanical pain threshold.

#### Temporal Summation of Pain (TSP)

was assessed using single stimulus in triplicate with a 256 mN pinprick (MRC Systems GmbH) applied to the skin surface of the selected sites, followed by a series of 10 identical stimuli (1 Hz – metronome-guided). TSP was calculated as the average pain rating from the series of 10 stimuli minus the average pain rating from the three trials with the single stimulus.

#### Pressure Pain Threshold (PPT) / Pressure Pain Tolerance (PPTol)

was assessed using a digital, handheld, clinical grade pressure algometer (Algometer II, Somedic SenseLab AB, Sweden). The pressure was manually increased at a rate of 50 kPa/s (1000 kPa max) until participants indicated that the sensation of pressure becomes one of faint pain (PPT) and the maximum pressure pain that the participant can tolerate (PPTol), respectively. The average of 3 trials per site was used for analysis.

#### Conditioned Pain Modulation (CPM)

A sustained pressure pain stimulus was used as the conditioning stimulus delivered using a cuff inflator on the gastrocnemius muscle of the non-dominant leg to elicit moderate pain (pain rating at 40-60 on a scale of 100) (35, 36). The PPT was then measured 3 times at the primary site and at the dominant trapezius muscle prior to and during the cuff stimulation. Pain ratings were obtained every 15 s prior to and during cuff conditioned stimuli. CPM magnitude was calculated as PPT during cuff pressure minus PPT at baseline .

### 2.5. SCD specific outcomes and morphine milligram equivalent (MME)

The severity and frequency of pain crises were assessed by the Adult Sickle Cell Quality-of-Life Measurement Information System (ASCQ-ME) (37), and the number of patient-reported pain crises in the preceding 12 months was documented, a method which is commonly used in pain research for SCD (18, 38). Pain-related QoL was evaluated using the Pediatric Quality of Life Inventory (PedsQL) (39). The average MME in the preceding 12 months was calculated using prescription data for opioid use before study enrollment. These data were retrieved from patients’ medical records and used to calculate the mean daily MME for each opioid, following the Centers for Disease Control recommended MME dose calculation formula (40): {strength per unit X number of units X number of prescriptions X MME conversion factor} / number of days = MME/Day.

### 2.6. Hematological analysis

Laboratory blood testing, including complete blood cell count, reticulocytes, and hemoglobin electrophoresis, was conducted on the day of TCM diagnosis (Supplementary Table 5).

### 2.7. Statistical analysis

Assessment of differences between TCM syndrome groups was performed using one-way analysis of variance (ANOVA) followed by Tukey post hoc testing. Two-sample t-tests (two tailed) were used to compare a subset of SCD patents and matched HCs at baseline. P-values less than 0.05 were considered significant. All analyses were conducted using GraphPad Prism software.

## 3. Results

### 3.1. Demographics

Demographic information is shown in Table 1. TCM groups did not differ in age (p=0.373), although the Stagnation group presented with a lower median age as compared to other groups. The Deficiency group showed significantly lower weight and body mass index (BMI) as compared with both the HCs and Stagnation group (p=0.0032; p=0.0002), respectively. The Stagnation group also had more participants numerically with chronic transfusion compared to the Deficiency and Equal groups. The distribution of hydroxyurea users was also numerically higher in the Deficiency group compared to the other two groups, but this was not statistically significant. Each SCD genotype was equally distributed across the three TCM groups.

### 3.2. Clinical Characteristics

All three TCM syndrome groups showed increased pain intensity and interference, as well as symptoms of neuropathic pain, compared to the control group (Table 2). However, only the Stagnation and Deficiency groups reported worse sleep quality, increased levels of fatigue, anxiety, and depression relative to HCs. In addition, it is worth noting that patients with SCD also reported pain in more body areas on the Widespread Pain Index and higher overall fibromyalgia severity. In a subset of SCD patients and matched HCs, the patients reported comprehensive physiological and psychological dysfunction (Supplementary Table 3), consistent with previous studies (17, 20, 28, 37).

**Table 2.** TCM Groups Differed on Patient Reported Outcomes. Patient reported outcomes in related with pain (intensity and interference), neuropathic and nociplastic pain syndromes, physical function, sleep, and fatigue were assessed with PROMIS-29, PainDETECT, Fibromyalgia Survey Questionnaires, the Pittsburgh Sleep Quality Index, Multi-dimensional Fatigue Inventory, as well as Hospital Anxiety and Depression Scale. Patients with SCD reported increased pain, physiological and psychological dysfunction as compared with HCs. The Stagnation and Deficiency groups displayed worse outcomes than the Equal group.

| Patient-reported outcomes | Score name (value range) | TCM#1 (Equal)<br>(Mean ± SD) | TCM#2 (Deficiency)<br>(Mean ± SD) | TCM#3 (Stagnation)<br>(Mean ± SD) | Healthy<br>(Mean ± SD) |
| --- | --- | --- | --- | --- | --- |
| PainDETECT | Total score (7-47) | 18.92 ± 3.825 **** | 18.25 ± 8.095 **** | 22.47 ± 9.172 **** | 7.273 ± 0.8013 |
| PROMIS-29 | Pain Intensity (0-10) | 3.583 ± 2.314 **** | 5.6 ± 1.639 ****# | 5.75 ± 1.88 ****## | 0.5000 ± 1.244 |
| | Pain Interference (4-20) | 11.50 ± 5.072 **** | 16.63 ± 5.830 ****## | 17.27 ± 3.369 ****## | 4.781 ± 1.791 |
| | Physical function (4-20) | 6.417 ± 1.975 | 8.438 ± 3.949 *** | 10.00 ± 4.536 ****# | 4.333 ± 1.407 |
| | Satisfaction with social role (4-20) | 14.42 ± 5.368 | 7.875 ± 3.757 ****### | 9.933 ± 4.574 ****# | 17.50 ± 3.556 |
| Fibromyalgia Survey<br>Questionnaire | Widespread Pain Index (0-19) | 4.667 ± 4.228 ** | 7.563 ± 4.066 **** | 6.267 ± 4.496 **** | 0.6061 ± 1.391 |
| | Symptom Severity Score (0-12) | 2.917 ± 1.832 | 5.313 ± 3.516 **** | 6.667 ± 2.637 ****## | 1.606 ± 1.999 |
| | Total Score (0-31) | 7.583 ± 5.632 * | 12.88 ± 6.879 ****# | 12.67 ± 5.367 ****# | 2.212 ± 3.090 |
| Pittsburgh Sleep Quality<br>Index (PSQI) | Global PSQI Index | 12.17 ± 8.032 | 20.73 ± 9.293 ***# | 20.80 ± 8.046 ***# | 10.91 ± 6.760 |
| Multi-dimensional Fatigue<br>Inventory (MFI) | Total MFI Score (0-100) | 45.58 ± 9.395 | 56.00 ± 19.73 * | 61.60 ± 15.29 ***# | 41.91 ± 13.46 |
| Hospital Anxiety and<br>Depression Scale | Anxiety Score (0-21) | 4.250 ± 2.417 | 6.250 ± 3.924 * | 8.714 ± 5.413 ****# | 2.719 ± 2.679 |
| | Depression Score (0-21) | 2.833 ± 2.038 | 5.250 ± 3.697 ** | 6.714 ± 3.750 ****## | 1.938 ± 1.795 |

\***p**< 0.05 compared with Healthy, \*\***p**< 0.01 compared with Healthy, \*\*\***p**< 0.001 compared with Healthy, \*\*\*\***p**< 0.0001 compared with Healthy, #**p**< 0.05 compared with Equal, ##**p**< 0.01 compared with Equal, ###**p**< 0.001 compared with Equal.

Compared to the Equal group, both the Stagnation (p=0.0013) and Deficiency (p=0.0046) groups reported increased multi-dimensional pain interference. Both groups also showed higher total scores for the FSQ (p=0.0328 Vs Deficiency and p=0.0477 Vs Stagnation) compared to the Equal group, while the Stagnation group displayed significantly higher Symptom Severity Scores (p=0.0013). Additionally, both the Deficiency and Stagnation groups exhibited worse sleep quality than the Equal group (p=0.0293 Vs Deficiency and p=0.0277 Vs Stagnation). Furthermore, compared to the Equal group, the Stagnation group showed significantly reduced physical function (p=0.0131) and elevated fatigue (p=0.0342), anxiety (p=0.0119), and depression (p=0.0035).

Notably, the Stagnation (n=6.526 ± 4.718) and Deficiency (n=5.19 ± 3.855) groups exhibited a higher frequency of pain crises compared to the Equal group (n=1.714 ± 1.684) (Fig.1A) along with higher combined pain crisis recency/severity as assessed by the ASCQ-ME (Fig. 1C), and lowest SCD pain-related QoL (Fig 1E), despite no difference in pain severity during crisis between groups (Fig. 1D). Additionally, both Stagnation (p=0.0011) and Deficiency (p=0.0029) groups had significantly higher opioid usage over the past 12 months compared to the Equal group (Fig. 1B).

**Figure 1.**
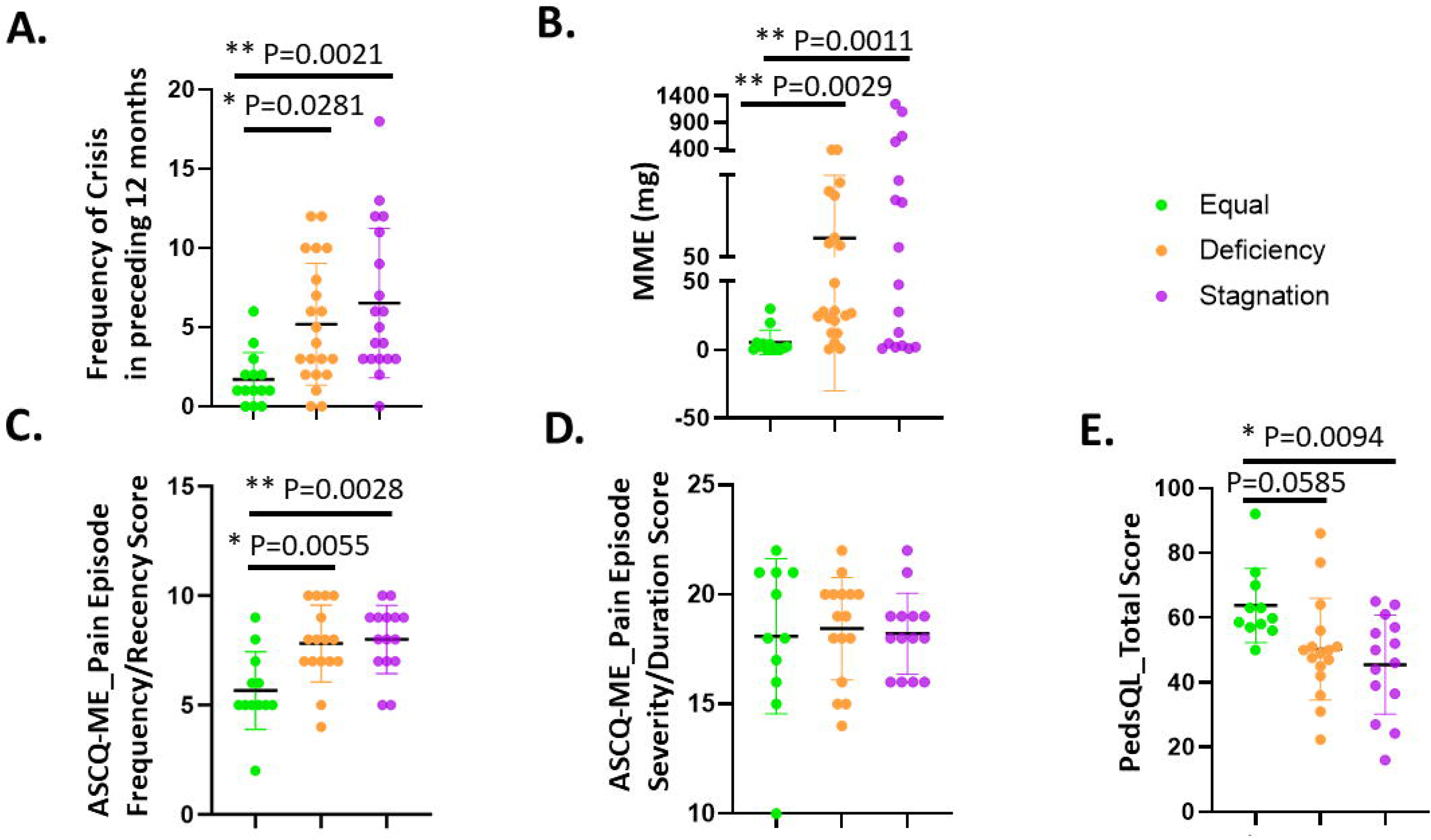
SCD Patients Differed on VOC, MME and QoL Across TCM Groups. The severity and frequency of pain crises and SCD Pain-related QoL were assessed by the ASCQ-ME (**C**) and PedsQL (**E**), respectively while the number of patient-reported pain crises in the preceding 12 months was also shown (**A**). The average MME in the preceding 12 months was calculated using: {strength per unit X number of units X number of prescriptions X MME conversion factor} / number of days = MME/Day. Stagnation and Deficiency groups exhibited a higher frequency of pain crises compared to the Equal group (n=1.714 ± 1.684) (**A and C)** and lowest SCD pain-related QoL (**E**). No difference in pain severity during crisis between groups (**D**). Importantly, both Stagnation and Deficiency groups had significantly higher opioid usage over the past 12 months compared to the Equal group (**B**).

#### QST

Interestingly, no significant differences between TCM groups on any QST outcomes were detected (Table 3), although Deficiency showed an insignificantly elevated MPT (95.38 ± 57.83) and HPT (8.940 ± 2.817) in the forearm whereas Equal group displayed a slightly increased PPT (243.9 ± 94.83) in the trapezius as compared with two other groups for each test. Consistent with previous studies, the differences between SCD and their matched healthy controls exhibited robust changes across mechanical and thermal sensitivity indexes (Supplementary Table 4). In a separate matched subset of SCD patients (n=28) and HCs (n=28), the patients exhibited decreased mechanical detection thresholds (p=0.0261 at the primary painful site), increased mechanical pain thresholds (primary painful site: p=0.0238; forearm: p=0.0358), pressure pain thresholds (primary painful site: p=0.0003; trapezius: p=0.0107), pressure pain tolerance (primary painful site: p=0.0009; trapezius: p = 0.0186), and cold pain thresholds (primary painful site: p=0.0332; forearm: p = 0.0250), indicating an overall pattern of increased mechanical and thermal sensitivity in SCD.

**Table 3.** TCM Groups Did not Differ on QST. . No significant differences between TCM groups on any QST outcomes were detected although Deficiency showed an insignificantly elevated Mechanical Pain Threshold and Heat Pain Threshold in the forearm whereas Equal group displayed a slightly but insignificantly increased Pressure Pain Threshold in the trapezius as compared with two other groups.

| **Score name** | **TCM#1 (Equal)**<br>**(Mean ± SD)** | **TCM#2 (Deficiency)**<br>**(Mean ± SD)** | **TCM#3 (Stagnation)**<br>**(Mean ± SD)** | **Healthy**<br>**(Mean ± SD)** |
| --- | --- | --- | --- | --- |
| Mechanical Detection Threshold<br>(forearm) | 0.2673 ± 0.1852 | 0.3346 ± 0.6821 | 0.4193 ± 0.2986 | 0.4276 ± 0.2605 |
| Mechanical Pain Threshold<br>(forearm) | 61.00 ± 61.01 | 95.38 ± 57.83 | 70.68 ± 49.89 | 93.64 ± 85.84 |
| Temporal Summation<br>(forearm) | 14.31 ± 11.94 | 12.46 ± 9.314 | 15.13 ± 12.70 | 11.54 ± 11.90 |
| Cold Detection Threshold<br>(forearm) | -1.983 ± 0.7309 | -1.947 ± 0.7643 | -2.907 ± 1.328 | -2.400 ± 1.147 |
| Cold pain Threshold<br>(forearm) | -8.864 ± 7.710 | -10.19 ± 7.673 | -9.147 ± 5.725 | -14.41 ± 9.501 |
| Heat Detection Threshold<br>(forearm) | 3.508 ± 1.179 | 3.233 ± 1.174 | 3.820 ± 2.474 | 3.424 ± 1.710 |
| Heat pain Threshold<br>(forearm) | 6.892 ± 4.024 | 8.940 ± 2.817 | 7.927 ± 3.499 | 8.038 ± 3.296 |
| Pressure Pain Threshold<br>(Trapezius) | 243.9 ± 94.83 | 215.1 ± 100.5 | 219.5 ± 76.08 | 271.8 ± 122.8 |
| Pressure Pain Tolerance<br>(Trapezius) | 336.9 ± 145.5 | 324.0 ± 124.8 | 386.6 ± 175.7 | 439.6 ± 244.0 |
| Conditioned Pain Modulation<br>(Trapezius) | 46.67 ± 56.90 | 49.21 ± 101.8 | 70.71 ± 105.2 | 35.53 ± 54.81 |

#### Hematological Indexes

Fresh blood samples from all patients were analyzed for CBC with differential and hemoglobin electrophoresis (refer to Supplementary Table 5). Most hematological indexes did not present any differences across the three different TCM syndrome groups. However, the Deficiency and Stagnation groups, but not the Equal group, exhibited significantly higher WBC levels compared to HCs.

## 4. Discussion

Pain and other symptoms are heterogeneously expressed across SCD patients (41). Indeed, the existing literature supports the notion that not all SCD patients have the same underlying pathophysiology, similar to other chronic pain disorders (42). The present work describes individual characteristics that differ between patients with SCD that are categorized by TCM diagnosis.

Chronic anemia, recurrent acute vaso-occlusive, and chronic pain are common manifestations of SCD caused by complex pathophysiology from hemoglobin S polymerization, hemolysis, endothelial dysfunction, and vaso-occlusion (43). According to the theory of TCM, symptoms such as low energy or excessive fatigue, sallow complexion, and pale lips and nails, which are caused by chronic anemia, are considered clinical manifestations of deficiency of both qi and blood (9). Recurrent pain in the chest, waist, back, and limbs, including severe pain and swelling of hands and feet caused by capillary microthrombosis, are considered clinical manifestation of blood stasis blocking meridians (44). As such, from a TCM perspective, the pathogenesis of SCD primarily manifests as a deficiency of both qi and blood, accompanied by blood stasis obstructing the meridians. Many individuals across our three TCM syndrome groups consistently displayed anemic manifestations such as pale lips and nails. The features of tongue observed in the TCM syndrome groups mainly reflected blood stasis-related manifestations, such as purple/dark spots in the tongue body. Many individuals across all TCM syndrome groups also presented with weak or thin wiry pulse, with more individuals in the Stagnation group who exhibited knotted pulse features indicating stagnation syndrome. These are consistent with characteristics of the “Bi Syndrome” that has insufficient righteousness and obstruction of meridians by pathogenic factors (45). Moreover, because of its severe pain as the signature of the condition, it may be considered as a form of “Painful Bi” and “Blood Stasis Bi” within the scope of “Bi Syndrome” as the intense pain is mainly caused by the obstruction of meridians by blood stasis.

Individuals with SCD enrolled in our study were classified into three main TCM syndromes: Equal, Deficiency, and Stagnation. These syndromes were primarily differentiated on the characteristics of pain (e.g., quality, intensity, onset, time of duration), physical dysfunction (e.g., fatigue/weak), followed by other symptoms (e.g., ease of sweating, facial complexion, voice, lips or nails, characteristics of “Tongue” and “Pulses”). See Supplementary Table 2 for a full description of diagnostic criteria.

The three TCM syndrome groups were differentiated in patient-reported outcomes that were related to pain and somatic symptoms, such as fatigue, physical function, sleep disturbance, as well as SCD related outcomes, including VOC specific pain and frequency and opioid consumption. Noticeably, the Stagnation and Deficiency groups showed greater nociplastic pain features and worse sleep compared to the Equal group. The Stagnation group also exhibited the lowest QOL scores compared to the Equal group. Additionally, the Stagnation group also displayed worse physical dysfunction, and higher levels of anxiety and depression as compared to the other two groups. These results suggest that the Stagnation group may have more nociplastic pain symptoms and other characteristics (sleep/fatigue/mood) that are commonly amplified in fibromyalgia, the canonical nociplastic pain condition. Interestingly, the three groups did not differ significantly in neuropathic pain or QST outcomes (see below). These data suggest that the three TCM groups did not differ with respect to nociceptive and neuropathic pain. Notably, both the Stagnation and Deficiency groups also had more intense pain-related clinical symptoms (Table 2) but differed in the duration and character of pain at the chronic stage of SCD and during acute SCD episodes (Fig.1).

With respect to SCD pathology, both Stagnation and Deficiency groups presented with higher frequency of VOCs (Fig. 1A and C) and opioid usage (MME) compared to Equal group (Fig. 1B). This increase in opioid requirement in the Stagnation group is consistent with nociplastic pain patients having reduced endogenous opioid efficacy (46). Importantly, the Stagnation group also reported the most frequent, intense, and sudden onset of VOCs as their main symptoms/signs, whereas the chief complaint of Deficiency group was more reflected in fatigue, weakness and chronic pain as summarized in Supplementary Table 2. This may explain the difference in general overall pain interference reported across the TCM syndrome groups, with the Stagnation group reporting the highest pain interference.

We also observed differences between the Deficiency and other two groups based on pain presentation. Noticeably, chronic pain was not always present and was sometimes absent in individuals of both Equal and Stagnation groups. The Deficiency group experienced more profound chronic pain along with Qi and Blood deficiency syndrome as reported in Supplementary Table 2. This long-term persistent pain with recurrent VOCs in the Deficiency group could be related to the significantly lower weight and BMI (Table 1) and RBC (Supplementary Table 5) as compared with Stagnation, which may further explain other co-existing symptoms such as intense fatigue, weakness, and profuse sweating that are also associated with “deficiency” in TCM (Supplementary Table 2).

The symptoms that may reflect a higher prevalence of “blood stasis” were more prominent in the Stagnation group whereas symptoms of more “blood and Qi Deficiency” were found in the Deficiency group from the TCM perspective. These data suggest a more personalized acupuncture treatment with selected acupoints and corresponding needling manipulations (“reinforce” / “reduce” / “non-reinforce or reduce”) may be needed to treat individual patients based on selected acupoints based on both TCM syndrome differentiation and individual symptoms.

Interestingly, the SS type of SCD seems to be equally distributed across the three TCM syndrome groups. The SCD subtype HbSS is known to be a more severe form of the disease on average compared to the HbSC genotype (47, 48), and it is positively correlated with the degree of psychological symptoms experienced by the patient (48, 49). More importantly, even though the WBC % was lower in the Equal group, this group presented with comparable levels of all pathological hematological indexes as compared to the two other groups (Supplementary Table 5). This finding may suggest that the less severe clinical symptoms presented in the individuals of Equal group are not related to the SCD genotype but may be associated with other underlying pathophysiological mechanisms in these patients.

In an effort to examine participant differences in touch and pain sensitivity we also conducted a battery of QST using methods that have been widely applied across various chronic pain conditions. Existing literature documenting altered QST thresholds using fixed testing sites in SCD has identified increased sensitivity in response to thermal/mechanical painful stimuli compared to non-SCD pain-free HCs (17, 33, 34, 50). However, pain in SCD can occur anywhere in the body or sometimes throughout the entire body during an active crisis, and the features and locations of pain can vary significantly among individuals. In our study, patients reported a variety of painful locations including: the lower- and mid- to upper-back, hips, shoulders, thighs, knees, and lower extremities. This occurred during both the steady phase and active crises. Moreover, the painful locations during acute pain episodes under VOC could also be exaggerated or replaced by more painful regions elsewhere (e.g., chest, large joints) or a combination of locations. We did not detect any significant differences in QST outcomes across the three TCM patterns. This suggests that our TCM syndromes were not different across nociceptive experimental pain stimuli. Of note, the 3 TCM groups also did not differ in neuropathic pain symptoms assessed with the PainDETECT questionnaire. As mentioned above, the Stagnation and Deficiency groups displayed higher nociplastic pain symptoms than the Equal groups which was driven mostly by the Symptom Severity sub-scale for the Stagnation group. Overall, these data may suggest involvement of nociplastic pain mechanisms in SCD moreso in the Stagnation and Deficiency groups than the Equal group.

Our study has limitations. This is a cross-sectional study; therefore, causation cannot be determined. For example, the two TCM patterns (“Stagnation” and “Deficiency”) had significantly higher opioid consumption (MME) than “Equal” pattern before enrollment, as such we could not determine if there was a relationship between the level of opioid consumption and TCM pattern. Moreover, we could not gauge the potential effects of this medication usage on TCM patterns.

The consistent performance of accurate diagnosis relies on the clinical experience of the practitioner and requires training. Our diagnosis criteria were also established based on the enrolled SCD patients in this study and therefore may not be a definitive reference for these results. In addition, the 3 TCM syndromes did not differentiate in other objective parameters such as laboratory exams or QST but primarily in patient-reported outcomes. Thus, this study lacks objective markers associated with different TCM patterns. Future studies should consider a more comprehensive diagnosis regimen at different stages of the disease (e.g., patients at active VOC stages can have different subtypes of Stagnation). The diagnosis criteria should be further validated and standardized in a larger sample size.

To our knowledge, this is the first and largest clinical trial investigating TCM syndrome diagnoses in SCD. Optimal therapeutic effects could be achieved by administering the TCM-guided treatment protocol with the appropriate acupuncture points and needling manipulations (51). The implementation of TCM syndrome diagnosis can not only guide the acupuncture treatment but also other alternative treatment such as herbology to facilitate consistent and personalized integrative care. The present study provides a novel and objective clinical relevance for TCM diagnosis and lays the foundation for assessing the outcomes of TCM-guided interventions for managing pain not only in SCD but also other clinical population with complex pathophysiology and large heterogeneity needing individualized treatment for optimal clinical outcomes. More in-depth investigations are warranted to enhance evidence-based integrative and complementary pain management in challenging clinical populations such as SCD.

## Data Availability

All data produced in the present study are available upon reasonable request to the authors.

## 5. Author Contributions

Y.W., D.D.W., and A.Q.P. analyzed, validated, and interpreted the data; Y.W. drafted the manuscript; D.D.W., S.E.H., and R.E.H. revised the manuscript and helped with data interpretation; Y.W. and A.R.O. directed patient recruitment; Y.W. designed the study, supervised the overall study performance, and edited the manuscript.

## 6. Acknowledgement

The authors would like to thank Brandon Alec Reyes, Tyler James Barret, Nayana Dutt, Payton Mittman, and Bea Paras for assisting with data collection; Candice Debats and Justin Smith for assisting with chart info retrieval and data summarization; and Siddhi Gandhi, Charanjit Kaur, and Veena Vijayan for data management and validation, as well as all the providers affiliated with Indiana University Health and Indiana Hemophilia & Thrombosis Center for patient referral to this study. The authors also thank Indiana University Clinical Research Center and Indiana University Research MRI Center for resource support.

---

**Supplementary Table 1.**
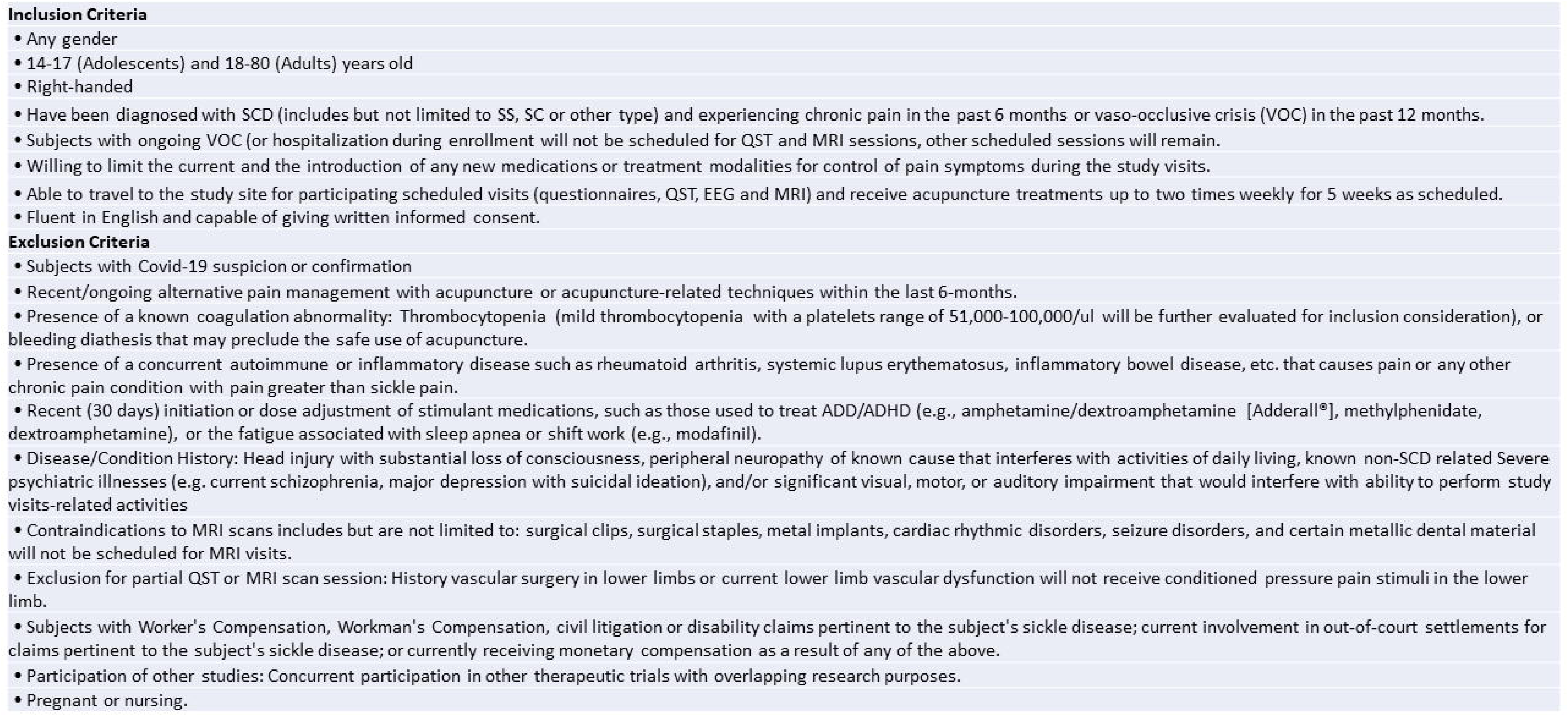
Study Eligibility Criteria.

**Supplementary Table 2.** TCM Diagnosis Criteria.

| TCM Groups<br>Diagnosis | TCM#1<br>Equal Deficiency and Stagnation | TCM#2<br>Deficiency Greater Than Stagnation | TCM#3<br>Stagnation Greater Than Deficiency |
| --- | --- | --- | --- |
| Main Symptoms/signs | Mild or moderate chronic pain only, or infrequent acute pain only, or chronic pain with infrequent acute pain<br>Tongue body: light red or slightly pale, or with purple spots<br>Pulses: thin, wiry, or normal | Group Symptoms 1:<br>Fatigue; weakness; shortness of breath; worsening after exertion; dull, sallow or pale complexion; pale lips and nails<br>Group Symptoms 2:<br>chronic pain only, or chronic pain that gets progressively worse before VOC; dull, or stabbing or sharp pain; mild or moderate or severe pain level; pain better by pressing<br>Tongue body: pale or pale with purple spots<br>Pulses: thin, weak and /or deep | Group symptoms 1:<br>Chronic pain with frequent sudden onset of acute pain; stabbing or sharp pain; moderate or severe pain level; pain aggravated by pressing<br>Group symptoms 2:<br>Fatigue; weakness; shortness of breath; worsening after exertion; dull, sallow or pale complexion; pale lips and nails<br>Tongue body: pale with purple spots, or dark<br>Pulses: wiry, or choppy or knotted |
| TCM diagnostic Criteria | Meet neither TCM#2 nor TCM#3 | Meet:<br>Four or more in group symptoms 1; two or more in group symptoms 2<br>Or<br>Three or more in group symptoms 1; two or more symptoms in group symptoms 2; Tongue and pulse signs | Meet:<br>Three or more symptoms in group symptoms 1; two or more in group symptoms 2<br>Or<br>Two or more in group symptoms 1; two or more in group symptoms 2; tongue and pulse signs |

**Supplementary Table 3.** Patient with SCD Differed with HCs in Patient Reported Outcomes.

| Patient-reported outcomes | Score name (value range) | Healthy (Mean ± SD) (n=28) | Matched SCD (Mean ± SD) (n=28) | P value |
| --- | --- | --- | --- | --- |
| PainDETECT | Total score (7-47) | 7.259 ± 0.8130 | 15.11 ± 10.51 | **<0.0007 \*\*\*** |
| PROMISE-29 | Pain Intensity (0-10) | 0.3846 ± 1.169 | 4.500 ± 2.232 | **<0.0001 \*\*\*\*** |
| | Pain Interference (4-20) | 4.731 ± 1.867 | 12.54 ± 7.201 | **<0.0001 \*\*\*\*** |
| | Physical function (4-20) | 4.333 ± 1.544 | 9.852 ± 4.312 | **<0.0001 \*\*\*\*** |
| | Satisfaction with social role (4-20) | 17.77 ± 3.278 | 8.885 ± 5.694 | **<0.0001 \*\*\*\*** |
| Fibromyalgia Survey Questionnaire | Widespread Pain Index (0-19) | 0.4444 ± 1.188 | 6.259 ± 3.996 | **<0.0001 \*\*\*\*** |
| | Symptom Severity Score (0-12) | 1.556 ± 1.928 | 5.296 ± 2.399 | **<0.0001 \*\*\*\*** |
| | Total Score (0-31) | 2.000 ± 2.746 | 9.333 ± 5.704 | **<0.0001 \*\*\*\*** |
| Pittsburgh Sleep Quality Index (PSQI) | Global PSQI Index (0-47) | 9.880 ± 5.681 | 20.52 ± 6.325 | **<0.0001 \*\*\*\*** |
| Multi-dimensional Fatigue Inventory | Total Score (0-100) | 40.85 ± 12.95 | 53.92 ± 17.90 | **0.0062 \*\*** |
| Hospital Anxiety and Depression Scale | Anxiety Score (0-21) | 2.680 ± 2.839 | 18.16 ± 19.38 | **0.0007 \*\*\*** |
| | Depression Score (0-21) | 2.040 ± 1.904 | 7.200 ± 6.069 | **0.0011 \*\*** |

**\*p< 0.05, \*\*p< 0.01, \*\*\*p< 0.001, \*\*\*\*p< 0.0001**

**Supplementary Table 4.** Patient with SCD Differed with HCs in QST.

| Score name | Matched Healthy<br>(n=28)<br>(Mean ± SD) | Matched SCD<br>(n=28)<br>(Mean ± SD) | P value |
| --- | --- | --- | --- |
| Mechanical Detection Threshold (Forearm) | 0.4435 ± 0.2664 | 0.3140 ± 0.2759 | 0.1152 |
| Mechanical Detection Threshold (Primary site) | 0.5368 ± 0.5209 | 0.2596 ± 0.2365 | **0.0261\*** |
| Mechanical Pain Threshold (Forearm) | 102.5 ± 91.86 | 58.56 ± 46.48 | **0.0358\*** |
| Mechanical Pain Threshold (Primary site) | 109.8 ± 64.27 | 66.37 ± 79.31 | **0.0238\*** |
| Mechanical Temporal Summation (Forearm) | 11.72 ± 12.22 | 12.30 ± 9.162 | 0.8571 |
| Mechanical Temporal Summation (Primary site) |  |  |  |
| Cold Detection Threshold (compared to BL skin temperature) (Forearm) | -2.396 ± 1.191 | -2.354 ± 1.124 | 0.9067 |
| Cold Detection Threshold (compared to BL skin temperature) (Primary site) | -2.723 ± 2.065 | -2.900 ± 1.724 | 0.7631 |
| Cold pain Threshold (compared to BL skin temperature) (Forearm) | -14.74 ± 9.513 | -9.082 ± 5.309 | **0.0250\*** |
| Cold pain Threshold (compared to BL skin temperature) (Primary site) | -15.26 ± 10.31 | -9.288 ± 6.539 | **0.0332\*** |
| Heat Detection Threshold (compared to BL skin temperature) (Forearm) | 3.589 ± 1.627 | 3.596 ± 1.654 | 0.9872 |
| Heat Detection Threshold (compared to BL skin temperature) (Primary site) | 4.233 ± 1.547 | 4.078 ± 2.103 | 0.8000 |
| Heat pain Threshold (compared to BL skin temperature) (Forearm) | 8.307 ± 3.234 | 8.126 ± 2.888 | 0.8534 |
| Heat pain Threshold (compared to BL skin temperature) (Primary site) | 8.522 ± 2.817 | 8.109 ± 2.805 | 0.6516 |
| Pressure Pain Threshold (Trapezius) | 296.0 ± 121.2 | 222.4 ± 79.85 | **0.0107\*** |
| Pressure Pain Threshold (Primary site) | 608.2 ± 304.2 | 372.3 ± 173.3 | **0.0003\*\*\*** |
| Pressure Pain Tolerance (Trapezius) | 466.8 ± 253.0 | 357.4 ± 177.4 | **0.0186\*** |
| Pressure Pain Tolerance (Primary site) | 362.9 ± 196.7 | 211.3 ± 92.31 | **0.0009\*\*\*** |
| Conditioned Pain Modulation (Trapezius) | 39.57 ± 61.13 | 50.11 ± 51.34 | 0.4759 |

\*p < 0.05, \*\*p < 0.01, \*\*\*p < 0.001

**Supplementary Table 5.**
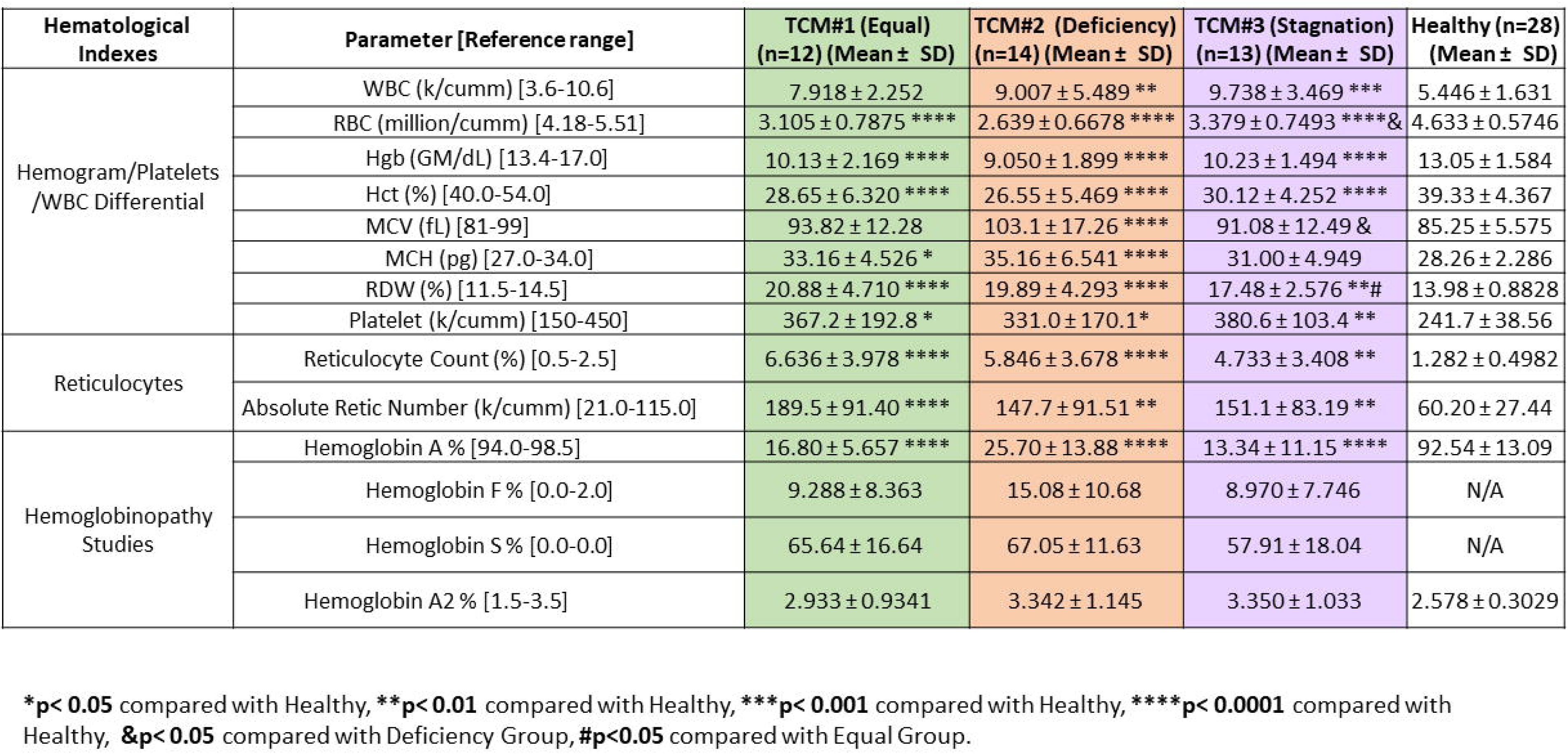
Patient with SCD Differed with HCs in Hematological Measures.

| Hematological Indexes | Parameter [Reference range] | TCM#1 (Equal)<br>(n=12) (Mean ± SD) | TCM#2 (Deficiency)<br>(n=14) (Mean ± SD) | TCM#3 (Stagnation)<br>(n=13) (Mean ± SD) | Healthy (n=28)<br>(Mean ± SD) |
| --- | --- | --- | --- | --- | --- |
| Hemogram/Platelets<br>/WBC Differential | WBC (k/cumm) [3.6-10.6] | 7.918 ± 2.252 | 9.007 ± 5.489 ** | 9.738 ± 3.469 *** | 5.446 ± 1.631 |
| | RBC (million/cumm) [4.18-5.51] | 3.105 ± 0.7875 **** | 2.639 ± 0.6678 **** | 3.379 ± 0.7493 ****& | 4.633 ± 0.5746 |
| | Hgb (GM/dL) [13.4-17.0] | 10.13 ± 2.169 **** | 9.050 ± 1.899 **** | 10.23 ± 1.494 **** | 13.05 ± 1.584 |
| | Hct (%) [40.0-54.0] | 28.65 ± 6.320 **** | 26.55 ± 5.469 **** | 30.12 ± 4.252 **** | 39.33 ± 4.367 |
| | MCV (fL) [81-99] | 93.82 ± 12.28 | 103.1 ± 17.26 **** | 91.08 ± 12.49 & | 85.25 ± 5.575 |
| | MCH (pg) [27.0-34.0] | 33.16 ± 4.526 * | 35.16 ± 6.541 **** | 31.00 ± 4.949 | 28.26 ± 2.286 |
| | RDW (%) [11.5-14.5] | 20.88 ± 4.710 **** | 19.89 ± 4.293 **** | 17.48 ± 2.576 **# | 13.98 ± 0.8828 |
| | Platelet (k/cumm) [150-450] | 367.2 ± 192.8 * | 331.0 ± 170.1* | 380.6 ± 103.4 ** | 241.7 ± 38.56 |
| Reticulocytes | Reticulocyte Count (%) [0.5-2.5] | 6.636 ± 3.978 **** | 5.846 ± 3.678 **** | 4.733 ± 3.408 ** | 1.282 ± 0.4982 |
| | Absolute Retic Number (k/cumm) [21.0-115.0] | 189.5 ± 91.40 **** | 147.7 ± 91.51 ** | 151.1 ± 83.19 ** | 60.20 ± 27.44 |
| Hemoglobinopathy<br>Studies | Hemoglobin A % [94.0-98.5] | 16.80 ± 5.657 **** | 25.70 ± 13.88 **** | 13.34 ± 11.15 **** | 92.54 ± 13.09 |
| | Hemoglobin F % [0.0-2.0] | 9.288 ± 8.363 | 15.08 ± 10.68 | 8.970 ± 7.746 | N/A |
| | Hemoglobin S % [0.0-0.0] | 65.64 ± 16.64 | 67.05 ± 11.63 | 57.91 ± 18.04 | N/A |
| | Hemoglobin A2 % [1.5-3.5] | 2.933 ± 0.9341 | 3.342 ± 1.145 | 3.350 ± 1.033 | 2.578 ± 0.3029 |

\***p**< 0.05 compared with Healthy, \*\***p**< 0.01 compared with Healthy, \*\*\***p**< 0.001 compared with Healthy, \*\*\*\***p**< 0.0001 compared with Healthy, &**p**< 0.05 compared with Deficiency Group, #**p**<0.05 compared with Equal Group.

